# MRI-derived lumbo-pelvic angles and lumbar discopathy in adults with low back pain from the Gaza Strip: a cross-sectional study

**DOI:** 10.64898/2025.12.29.25343004

**Authors:** Abdullah Abu Mousa, Yasser Alajerami, Ahmed Najim, Fahad Alghamdi, Kinan Mokbel

## Abstract

**Objective:** To examine the association between lumbo-pelvic angles (LPAs) and Magnetic Resonance Imaging (MRI) detected discopathy in adults with low back pain (LBP) in the Gaza Strip and to establish local reference values for LPA measurements.

**Methodology:** Prospective cross-sectional study, 200 adults with LBP referred for lumbosacral MRI at two major hospitals in Gaza Strip. 1.5T MRI scanners were used, and Lumbar lordosis angle (LLA), sacral kyphosis angle (SKA) and sacral table angle (STA) were measured on mid-sagittal T2 images. Discopathy characteristics were recorded, and disability was assessed using the Oswestry Disability Index.

**Result:** Of the 200 participants (mean age 45.7±13.6 years; 52.5% male), discopathy was most common at L4/L5 (89.5%), L5/S1 (67%) and L3/L4 (40.5%). LPAs were not significantly associated with discopathy involvement, type or severity, except for SKA discopathy severity at L3/4 (*p*=0.044). LPA measures were consistent across age groups, though LLA and STA were lower in males (*p*<0.001 and *p*=0.008), and obese individuals had higher LLA than those of normal weight (*p*=0.004). Reference LPA values were established stratified by LBP duration, in acute, subacute and chronic LBP, indicating a negative correlation between LLA and SKA in moderate and chronic duration.

**Conclusion:** In adults with LBP in Gaza Strip, MRI-derived LPA showed limited association with lumbar discopathy characteristics, pain duration or disability. Although small differences related to gender, BMI and a single disc level were observed, overall associations were weak. The study establishes population-specific reference values for LPAs, which should be interpreted cautiously within a broader clinical context.

## Introduction

Low back pain (LBP) is a highly prevalent manifestation, affecting the majority of adults at some point in their lives [1, 2]. Intervertebral disc degeneration is a key contributor to LBP and is strongly associated with ageing, with degenerative changes emerging in early adulthood and increasing with age [3–5]. The condition is a leading cause of activity limitation, work-related disability, and substantial healthcare and socioeconomic burden worldwide [1, 6]. Despite this impact, there remains no consensus on the relative contributions of anatomical, degenerative, and functional factors to LBP, and the clinical relevance of lumbo-pelvic morphology remains controversial, with inconsistent findings reported across imaging and clinical studies [7–9].

Degenerative spinal changes are highly prevalent, even among asymptomatic individuals [10]. The association between LBP and sagittal lumbopelvic alignment remains inconsistent, with LBP reported in relation to both increased and decreased lumbar lordosis and sacral inclination [11–14]. Several radiographic studies have demonstrated correlations between LBP and lumbosacral parameters, including Lumbar lordosis angle (LLA), sacral kyphosis angle (SKA) and sacral table angle (STA) [15–17]. Alterations in sagittal spinal shape have long been implicated in the development and persistence of LBP, with clinical and biomechanical evidence linking abnormal posture and spinopelvic parameters to chronic symptoms [18–20]. Although LBP is frequently attributed to degenerative disc disease, disc herniation, or spinal stenosis, and commonly investigated using Magnetic Resonance Imaging (MRI) or computed tomography (CT), imaging findings often show degenerative abnormalities of uncertain clinical relevance, with the majority of spine MRI scans in LBP patients demonstrating incidental degenerative changes [4, 5, 21, 22].

Multiple factors, including age, sex, obesity, smoking, socioeconomic status, occupational loading, inactivity, trauma, vibration exposure, and genetic predisposition, influence lumbar disc degeneration [23–26]. Although numerous studies have explored associations between lumbosacral morphology and degenerative disc disease, the nature and clinical relevance of these relationships remain unclear, indicating a need for further investigation [24, 26–29].

The lumbopelvic angle (LPA) approach may provide a practical surrogate framework for understanding and managing LBP, with potential implications for reducing disease burden and healthcare costs. Accordingly, this study aimed to evaluate the association between LPA and MRI-identified disc abnormalities in adults with LBP, examine correlations with pain duration and disability, assess variation in LPA across disc characteristics, demographics, and BMI, and establish local reference values for LPA measurements in discopathy-related LBP duration.

## Methods and Materials

### Study Design and Ethics Approval

A prospective cross-sectional study comprises patients with LBP referred for lumbosacral MRI at Al-Shifa Medical Complex and European Gaza Hospital (EGH), both governmental hospitals supervised by the Ministry of Health (MoH) in the Gaza Strip. Data was collected between June 2020 and January 2021. Ethical approval was granted by the Helsinki Committee (HC) for Ethical Approval, Palestinian Health Research Council (PHRC), Gaza Strip, Palestine (approval number PHRC/HC/713/20; 01 June 2020). Administrative approval was obtained from the Ministry of Health, State of Palestine (Human Resources Development Directorate; correspondence number 504832; 16 June 2020). Academic approval was granted by the Faculty of Applied Medical Sciences, Al-Azhar University-Gaza (03 June 2020). Written informed consent was obtained from all participants prior to participation. A consent form was obtained from participants, and the pilot study was conducted before data collection to assess feasibility and refine data collection procedures [Supplementary S1].

### Sample size

During the COVID-19 pandemic, elective lumbosacral spine (LSS) MRI examinations decreased to approximately 380 per month. Using the SurveyMonkey sample size calculator, a sample of 192 participants was estimated to provide 95% confidence at a 5% margin of error. To account for potential withdrawals and missing data, an additional eight cases were added, yielding a final target sample size of 200 participants.

### Study Population

Consecutive adult patients presenting with LBP and referred for LSS MRI at the two hospitals were screened for eligibility. Suspected LBP cases based on clinical complaints were enrolled after applying the inclusion and exclusion criteria:

Inclusion criteria:

- Adults aged 18–80 years.
- Referral for LSS MRI due to LBP, with or without radicular pain.
- Ability and willingness to provide informed consent.

Exclusion criteria:

- History of spinal malignancy or spinal cord tumour.
- Congenital lumbosacral anomalies (e.g. scoliosis, lumbarisation, sacralisation).
- History of vertebral fracture or prior spinal surgery.
- Pregnancy.

### Data Collection and Assessments

#### Magnetic Resonance Imaging (MRI)

MRI scanners at governmental hospitals in the Gaza Strip. Al-Shifa Medical Complex hospital has a Siemens Magnetom Aera 1.5 T MRI scanner (Siemens Healthineers AG, Siemensstr. 3, 91301 Forchheim, Germany), and EGH has a 1.5 T Philips Intera MRI scanner (Philips Medical Systems, Best, The Netherlands). MRI LSS under routine protocols, including T2WI-TSE, T1WI-TSE sagittal, and T2WI-TSE axial. To view sample data, extract diagnostic information, and measure LPAs via RadiAnt DICOM by Medixant (Promienista 25, 60-288 Poznań, Poland) was used.

#### Diagnostic information

MRI images were reviewed and reported in the diagnostic findings form [Supplementary S2] by four expert consultant MRI radiologists at both hospitals. These reports involve detailed identification of the involvement, types, side, location, and severity/size of intervertebral discopathy. Furthermore, Spinal canal stenosis, spondylolisthesis, retrolisthesis, Schmorl’s nodes, and Modic changes in the vertebral endplates were evaluated.

#### Measure lumbo-pelvic angles

LLA, SKA and STA were measured separately on the mid-sagittal T2WI plane, as they sufficiently express the lumbar and pelvic morphometry [16, 30]. The LLA is determined by lines passing through the midpoints of the L1 and L5 vertebrae. The STA is measured between the upper and posterior surfaces of the sacral endplate

#### Questionnaire

Sociodemographic and economic status: along with the Status of diseases and medical history, such as LBP history, types, disability measurement index, and previous treatments. The Oswestry Disability Index (ODI) questionnaire is used to measure disability in LBP and is considered the gold standard for low back functional outcomes [31]. It covers 10 topics: pain intensity, lifting, ability to care for oneself, ability to walk, ability to sit, sexual function, ability to stand, social life, sleep quality, and ability to travel. Each category is followed by six statements describing various scenarios in the patient’s life related to that topic. Each question is scored on a scale of 0 to 5, where the first statement scores 0 (indicating the least disability) and the last scores 5 (indicating the most severe disability). The scores for all answered questions are summed up and then multiplied by 2 to generate the index (range 0 to 100). Zero indicates no disability, and 100 represents maximum disability [32, 33]. Scoring and interpretation of each of the ten items in the ODI are as follows [32]:

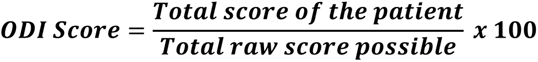

The total raw score is 50, and the patient’s total score depends on their answers.

- 0% –20%: Minimal disability
- 21%–40%: Moderate Disability
- 41%–60%: Severe Disability
- 61%–80%: Crippling back pain

- 81%–100%: Bed-bound.

### Statistical Analysis

The researcher used the Statistical Package for Social Sciences (SPSS Inc., version 25, Chicago, Illinois, USA) for data entry and statistical analysis. The quantitative data were analysed based on descriptive and inferential approaches. The central tendency parameters are also used (i.e. mean and standard deviation). The Kolmogorov-Smirnov test is used to assess the normality of each histogram, the T-test is used to test the study’s hypothesis as an independent T-test, and the ANOVA test is used to determine the significance of predisposing factors on LPA. Pearson’s correlation is used to investigate the correlation between LPA and LBP. The confidence interval was considered at 95% and had a margin of error of 5%. A *p*-value less than 0.05 was considered to indicate a statistically significant difference.

## Results

### Demographic and Characteristics

The participants’ characteristics are summarised in Table 1. The study involved 200 individuals with an average age of 45.7 years (±13.6). The largest group was middle-aged, making up 37%, followed by young adults at 36.5%. Males comprised 52.5% of the sample, while females made up 47.5%. By BMI, 42% were obese, 29.5% had normal weight, and 28.5% were overweight. Education was mainly secondary school (34%) or bachelor’s degree (28%), with 1.5% postgraduate. Lifestyle hazards linked to discopathy: long-standing (49.5%) and sitting (21%). The mean family income was 1200 ± 995.9.

**Table 1:** Distribution of the study participants by their demographic, clinical history variables, and Characteristics of clinical status for discopathy in the study population (n=200)

| Variables | (N) | (%) |
| --- | --- | --- |
| **Demographic** |  |  |
| **Age groups:** |  |  |
| 18-40 years | 73 | 36.5 |
| 41-55 years | 74 | 37 |
| 56-80 years | 53 | 26.5 |
| **Gender:** |  |  |
| Male | 105 | 52.5 |
| Female | 95 | 47.5 |
| **Body mass index:** |  |  |
| Normal weight | 59 | 29.5 |
| Overweight | 57 | 28.5 |
| Obese | 84 | 42 |
| **Education level:** |  |  |
| Elementary School | 34 | 17 |
| Secondary School | 68 | 34 |
| Diploma | 39 | 19.5 |
| Bachelor | 56 | 28 |
| Postgraduate | 3 | 1.5 |
| **Lifestyle hazards:** |  |  |
| No hazard | 21 | 10.5 |
| Hard work | 24 | 12 |
| Bending and lifting | 14 | 7 |
| Pre-long standing | 99 | 49.5 |
| Pre-long sitting | 42 | 21 |
| Monthly family income | Mean±(SD) | 1200 ± (995.9) |
| **Clinical History** |  |  |
| **History of chronic disease:** |  |  |
| No disease | 123 | 61.5 |
| Diabetes (DM) | 14 | 7.0 |
| Hypertension (HTN) | 33 | 16.5 |
| Cardiovascular disease (CVD) | 2 | 1.0 |
| Osteoarthritis (OA) | 3 | 1.5 |
| DM+HTN+CVD | 17 | 8.5 |
| HTN+OA | 1 | 0.5 |
| Lung disease | 1 | 0.5 |
| Endocrine disease | 3 | 1.5 |
| Others | 3 | 1.5 |
| **History of trauma:** |  |  |
| yes | 52 | 26.0 |
| No | 148 | 74.0 |
| **Previous imaging:** |  |  |
| No | 29 | 14.5 |
| X-ray | 79 | 39.5 |
| CT | 22 | 11.0 |
| MRI | 70 | 35.0 |
| **Discopathy family history:** |  |  |
| Yes | 98 | 49.0 |
| No | 102 | 51.0 |
| **Previous diagnosis:** |  |  |
| Yes | 102 | 51.0 |
| No | 98 | 49.0 |
| **Associated with cervical disc:** |  |  |
| Yes | 101 | 50.5 |
| No | 99 | 49.5 |
| **Take medication:** |  |  |
| Yes | 174 | 87.0 |
| No | 26 | 13.0 |
| **Medication response:** |  |  |
| Yes | 132 | 66.0 |
| No | 42 | 21.0 |
| No medication | 26 | 13.0 |
| **Pain after physiotherapy:** |  |  |
| Not change | 36 | 18.0 |
| Increase | 11 | 5.5 |
| Decrease | 40 | 20.0 |
| No physiotherapy | 113 | 56.5 |
| **Clinical Status for Discopathy** |  |  |
| **Discopathy complaint:** |  |  |
| LBP only | 18 | 9.0 |
| Radiated pain only | 16 | 8.0 |
| LBP with Radiated pain | 166 | 83.0 |
| **Radiculopathy sides:** |  |  |
| No radical pain | 18 | 9.0 |
| Right side | 46 | 23 |
| Left side | 72 | 36.0 |
| Bilateral side | 64 | 32.0 |
| **Radiculopathy symptoms:** |  |  |
| Asymptomatic | 24 | 12.0 |
| Numbness | 91 | 45.5 |
| Lower limb weakness | 5 | 2.5 |
| Numbness +weakness | 80 | 40.0 |
| **Walking difficulty:** |  |  |
| Yes | 141 | 70.5 |
| No | 59 | 29.5 |

The clinical history indicated that most participants reported no chronic disease (61.5%), and half (51%) reported no family history, while 74% had experienced a previous trauma. At the same time, 51% had a prior diagnosis of lumbar-only issues and combined lumbar-cervical discopathy was reported in 49.5% and 50.5%, respectively. Most used Nonsteroidal Anti-Inflammatory Drugs (NSAIDs) or muscle relaxants (87%); 66% reported benefit, and 21% no response. Physiotherapy was viewed skeptically by 56.5%; 20% reported reduced LBP, 18% no change, and 5.5% worsening.

Clinically, 83% of LBP cases were reported with radiating pain, while 9% were reported with LBP and 8% with radiated pain. Unilateral radiculopathy was more frequent on the left (36%) than on the right (23%), and 32% reported bilateral radiculopathy. Numbness (45.5%) was the most frequently reported symptom, followed by combined numbness and lower-limb weakness (40%). Lower-limb weakness alone was reported infrequently (2.5%). Walking difficulty was reported by 70.5% of participants, while 29.5% reported no walking difficulty.

### Characteristics of LBP for Discopathy Patients

Most participants reported chronic LBP, with 77% experiencing symptoms lasting longer than 12 weeks. Acute LBP (<6 weeks) was reported by 26%, while subacute LBP (6 to 12 weeks) was reported by 20%, as shown in Supplementary S3. Disability severity, assessed using the ODI, is presented in Supplementary S4. The largest group of participants was classified as having moderate disability (42.5%), followed by severe disability (34.5%). Minimal disability was observed in 13%, while crippling disability was reported in 10%.

### Presentation of MRI Findings for Discopathy Patients

The MRI findings for discopathy, summarised in [Table 2]. Discopathy was most common at L4/5 (89.5%), followed by L5/S1 (67%) and L3/4 (40.5%), with lower rates at L2/3 (13.5%) and L1/2 (5%). Disc bulge predominated at L4/5 (70%), L5/S1 (45%), and L3/4 (31.5%), while herniation and protrusion were less frequent (herniation: L5/S1 9.5%, L4/5 8.5%, L3/4 2.5%; and protrusion: L5/S1 9.5%, L3/4 6%, L4/5 5%), and caudal migration exceeded cephalic (peak 4.5% at L4/5 vs 1.5%).

**Table 2.** Distribution of MRI radiological findings by disc level for discopathy patients (n=200). N: number of participants; %: percentages.

| Variables | L1/2 |  | L2/3 |  | L3/4 |  | L4/5 |  | L5/S1 |  |
| --- | --- | --- | --- | --- | --- | --- | --- | --- | --- | --- |
|  | (N) | (%) | (N) | (%) | (N) | (%) | (N) | (%) | (N) | (%) |
| **Discopathy involvement:** |  |  |  |  |  |  |  |  |  |  |
| Yes | 10 | 5.0 | 27 | 13.5 | 81 | 40.5 | 179 | 89.5 | 134 | 67.0 |
| No | 190 | 95.0 | 173 | 86.5 | 119 | 59.5 | 21 | 10.5 | 66 | 33.0 |
| **Discopathy types:** |  |  |  |  |  |  |  |  |  |  |
| No Discopathy | 190 | 95.0 | 173 | 86.5 | 119 | 59.5 | 21 | 10.5 | 66 | 33.0 |
| Disc protrusion | 1 | 0.5 | 2 | 1.0 | 12 | 6.0 | 10 | 5.0 | 19 | 9.5 |
| Disc herniation | 2 | 1.0 | 1 | 0.5 | 5 | 2.5 | 17 | 8.5 | 19 | 9.5 |
| Disc bulge | 4 | 2.0 | 22 | 11.0 | 63 | 31.5 | 140 | 70.0 | 90 | 45.0 |
| Disc cephalic migration | 1 | 0.5 | 0 | 0.0 | 1 | 0.5 | 3 | 1.5 | 2 | 1.0 |
| Disc caudal migration | 2 | 1.0 | 1 | 0.5 | 0 | 0.0 | 9 | 4.5 | 3 | 1.5 |
| Disc sequestration | 0 | 0.0 | 1 | 0.5 | 0 | 0.0 | 0 | 0.0 | 1 | 0.5 |
| **Discopathy affected side:** |  |  |  |  |  |  |  |  |  |  |
| No disc | 190 | 95.0 | 173 | 86.5 | 119 | 59.5 | 21 | 10.5 | 66 | 33.0 |
| Right side/Asymmetric | 5 | 2.5 | 5 | 2.5 | 19 | 9.5 | 29 | 14.5 | 30 | 15.0 |
| Left side/Asymmetric | 1 | 0.5 | 9 | 4.5 | 16 | 8.0 | 58 | 29.0 | 39 | 19.5 |
| Both sides/diffuse | 1 | 0.5 | 12 | 6.0 | 39 | 19.5 | 85 | 42.5 | 35 | 17.5 |
| Central/Focal | 3 | 1.5 | 1 | 0.5 | 7 | 3.5 | 7 | 3.5 | 30 | 15.0 |
| **Discopathy location:** |  |  |  |  |  |  |  |  |  |  |
| No disc | 190 | 95.0 | 173 | 86.5 | 119.0 | 59.5 | 21.0 | 10.5 | 66 | 33.0 |
| Central zone | 3 | 1.5 | 1 | 0.5 | 8.0 | 4.0 | 10.0 | 5.0 | 29 | 14.5 |
| Subarticular zone | 3 | 1.5 | 2 | 1.0 | 5.0 | 2.5 | 16.0 | 8.0 | 18 | 9.0 |
| Foramina | 0 | 0.0 | 9 | 4.5 | 35.0 | 17.5 | 105.0 | 52.5 | 47 | 23.5 |
| Extra-foramina | 4 | 2.0 | 15 | 7.5 | 33.0 | 16.5 | 48.0 | 24.0 | 40 | 20.0 |
| **Discopathy size /severity:** |  |  |  |  |  |  |  |  |  |  |
| **Normal** | 190 | 95.0 | 173 | 86.5 | 119 | 59.5 | 21 | 10.5 | 66 | 33.0 |
| **Grade 1 (Mild)** | 4 | 2.0 | 13 | 6.5 | 39 | 19.5 | 78 | 39.0 | 67 | 33.5 |
| **Grade 2 (Moderate)** | 2 | 1.0 | 9 | 4.5 | 26 | 13.0 | 72 | 36.0 | 46 | 23.0 |
| **Grade 3 (Severe)** | 4 | 2.0 | 5 | 2.5 | 16 | 8.0 | 29 | 14.5 | 21 | 10.5 |
| **Other findings** |  |  |  |  |  |  |  |  |  |  |
| **Spinal canal stenosis:** |  |  |  |  |  |  |  |  |  |  |
| **Yes** | 10 | 5 | 3 | 1.5 | 20 | 10 | 20 | 10 | 30 | 15 |
| **No** | 190 | 95 | 197 | 98.5 | 180 | 90 | 180 | 90 | 170 | 85 |
| **Spondylolesthesis:** |  |  |  |  |  |  |  |  |  |  |
| **Yes** | 0 | 0 | 1 | 0.5 | 1 | 0.5 | 3 | 1.5 | 5 | 2.5 |
| **No** | 200 | 100 | 199 | 99.5 | 199 | 99.5 | 197 | 98.5 | 195 | 97.5 |
| **Retrolisthesis:** |  |  |  |  |  |  |  |  |  |  |
| **Yes** | 0 | 0 | 0 | 0 | 0 | 0 | 2 | 1 | 2 | 1 |
| **No** | 200 | 100 | 200 | 100 | 200 | 100 | 198 | 99 | 198 | 99 |
| **Schmorl's nodes:** |  |  |  |  |  |  |  |  |  |  |
| **Yes** | 37 | 18.5 | 36 | 18 | 36 | 18 | 23 | 11.5 | 7 | 3.5 |
| **No** | 163 | 81.5 | 164 | 82 | 164 | 82 | 177 | 88.5 | 193 | 96.5 |
| **Modic changes endplate:** |  |  |  |  |  |  |  |  |  |  |
| **No change** | 166 | 83 | 158 | 79 | 154 | 77 | 136 | 68 | 130 | 65 |
| **Type1 (bone marrow edema)** | 3 | 1.5 | 5 | 2.5 | 3 | 1.5 | 7 | 3.5 | 4 | 2 |
| **Type 2 (fatty change)** | 30 | 15 | 37 | 18.5 | 39 | 19.5 | 56 | 28 | 63 | 31.5 |
| **Type 3 (Sclerotic)** | 1 | 0.5 | 0 | 0 | 4 | 2 | 1 | 0.5 | 3 | 1.5 |

Regarding the discopathy-affected side, the nerve root compression was most frequent at L4/5 (42.5%). Compression is more asymmetric on the left side than the right at L3/4, L4/5, and L5/S1, with 8%, 29%, and 19.5% respectively. Most cases involve nerve compression in the foraminal region, mainly at L4/5 (52.5%) compared to extra-foraminal areas (24%), with mild discopathy. Other findings included stenosis (highest at L5/S1, 15%), low rates of spondylolisthesis and retrolisthesis, with Schmorl’s nodes mainly at the upper levels (L1/2, 18.5%; L2/3 and L3/4, 18%), and predominantly Modic type 2 changes (L5/S1, 31.5%; L4/5, 28%).

### The Correlation Between Lumbo-Pelvic Morphometric and LBP Duration, Disability Scores Based on MRI

The Pearson correlation between LPA and LBP characteristics, including LBP duration (yearly) and ODI disability scores summaries in Table 3. There was no statistically significant correlation between the LLA and LBP duration (*p*=0.518) or between LLA and ODI scores (*p*=0.47). Similarly, the SKA showed no statistically significant correlation with LBP duration (*p*=0.93) or ODI scores (*p*=0.57). The STA was also not significantly correlated with ODI scores (*p*=0.63). In contrast, a weak but statistically significant positive correlation was observed between STA and LBP duration (*r*=0.170, *p*=0.016).

**Table 3.** Distribution of Pearson Correlation between MRI LPA with LBP characteristics (n=200). LBP: lower back pain; LLA: Lumbar Lordosis Angle; SKA: Sacral Kyphosis Angle; STA: Sacral Table Angle; * Correlation is significant.

| LBP characteristics | LLA |  | SKA |  | STA |  |
| --- | --- | --- | --- | --- | --- | --- |
|  | *r* | *p* | *r* | *p* | *r* | *p* |
| **LBP duration yearly** | 0.046 | 0.518 | -0.005 | 0.938 | 0.170* | 0.016 |
| **ODI score** | 0.051 | 0.47 | 0.04 | 0.57 | -0.033 | 0.638 |

### The Relationship between lumbo-pelvic angles and Discopathy Involved, Types and Severity

The relationship between discopathy involvement, discopathy types (protrusion, herniation, bulge, cephalic/caudal migration, and sequestrated disc), and discopathy severity (normal, mild, moderate, and severe) with lumbo-pelvic morphometric (LLA, SKA, and STA) at the last three intervertebral lumbosacral disc spaces are shown in Table 4.

**Table 4.**
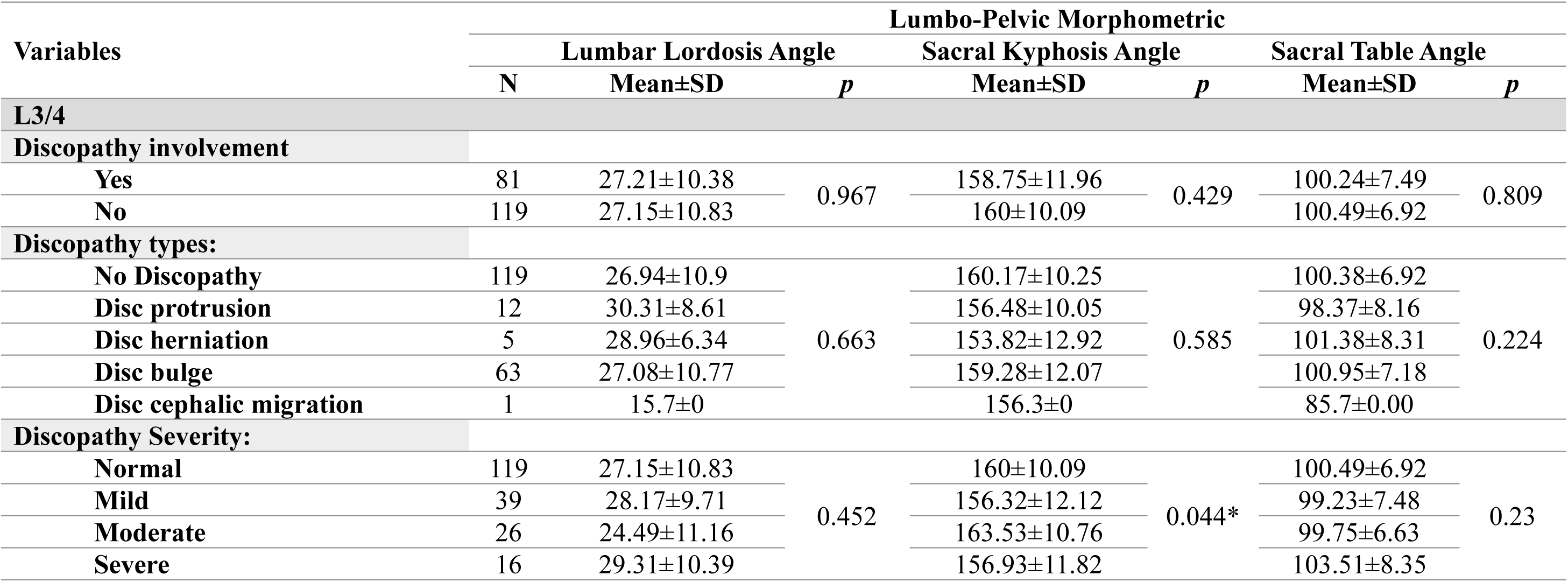
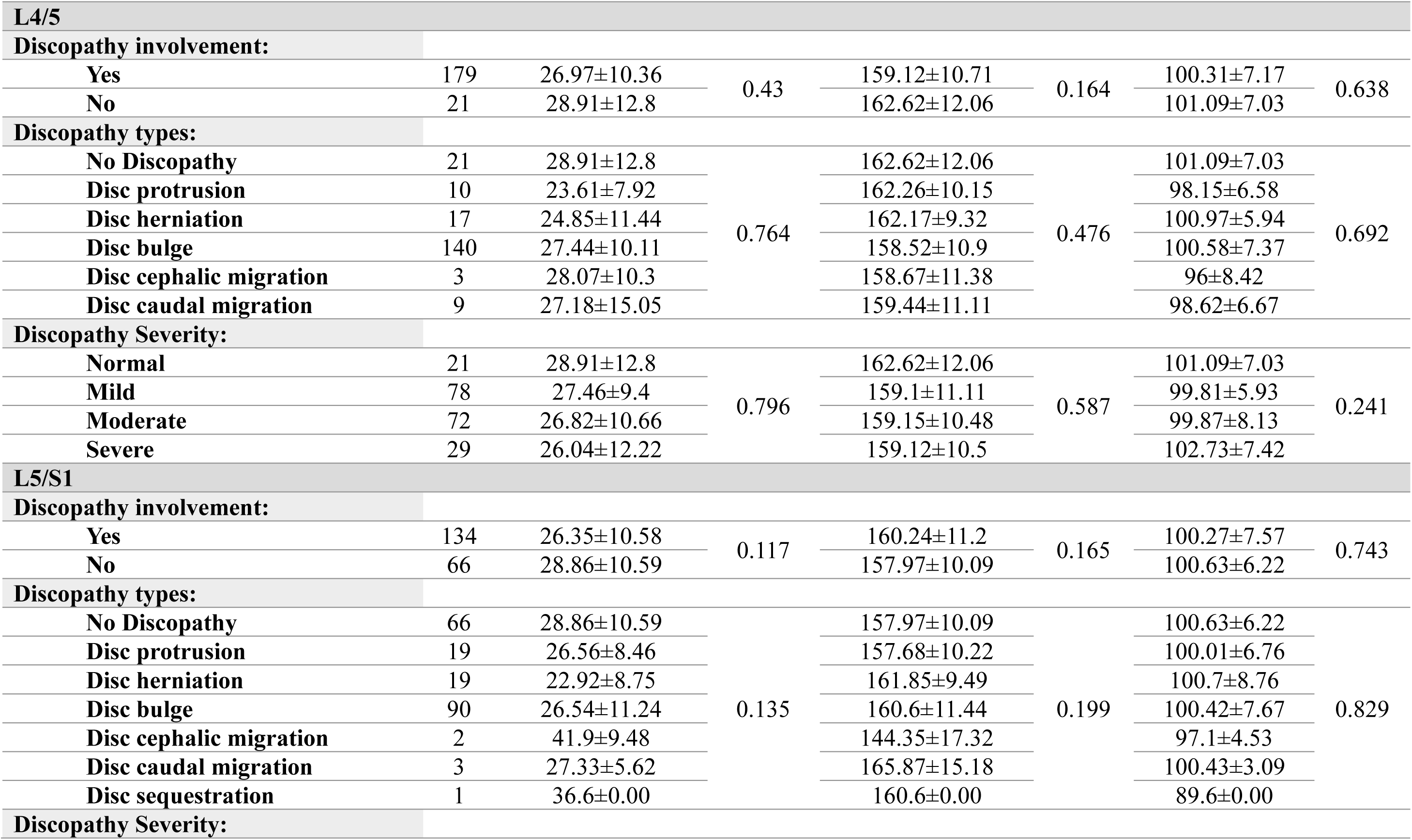
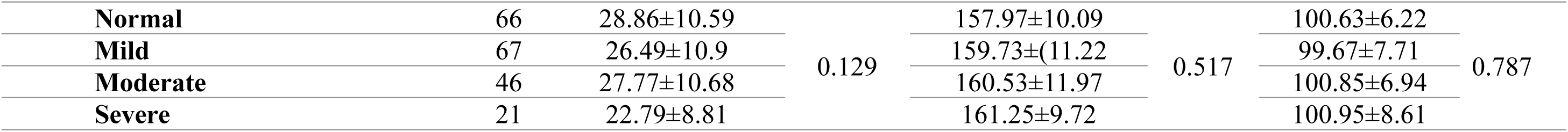
Distribution of the relationship between lumbo-pelvic morphometric (LPA) changes with discopathy involvement, discopathy types, and discopathy severity (n=200). N: number of participants; L: lumbar spine vertebra; S: sacral vertebra; SD: standard deviation; * relationship is significant

| Variables | Lumbo-Pelvic Morphometric |  |  |  |  |  |  |
| --- | --- | --- | --- | --- | --- | --- | --- |
|  | Lumbar Lordosis Angle |  |  | Sacral Kyphosis Angle |  | Sacral Table Angle |  |
|  | N | Mean±SD | *p* | Mean±SD | *p* | Mean±SD | *p* |
| **L3/4** |  |  |  |  |  |  |  |
| **Discopathy involvement** |  |  |  |  |  |  |  |
| Yes | 81 | 27.21±10.38 | 0.967 | 158.75±11.96 | 0.429 | 100.24±7.49 | 0.809 |
| No | 119 | 27.15±10.83 |  | 160±10.09 |  | 100.49±6.92 |  |
| **Discopathy types:** |  |  |  |  |  |  |  |
| No Discopathy | 119 | 26.94±10.9 | 0.663 | 160.17±10.25 | 0.585 | 100.38±6.92 | 0.224 |
| Disc protrusion | 12 | 30.31±8.61 |  | 156.48±10.05 |  | 98.37±8.16 |  |
| Disc herniation | 5 | 28.96±6.34 |  | 153.82±12.92 |  | 101.38±8.31 |  |
| Disc bulge | 63 | 27.08±10.77 |  | 159.28±12.07 |  | 100.95±7.18 |  |
| Disc cephalic migration | 1 | 15.7±0 |  | 156.3±0 |  | 85.7±0.00 |  |
| **Discopathy Severity:** |  |  |  |  |  |  |  |
| Normal | 119 | 27.15±10.83 | 0.452 | 160±10.09 | 0.044* | 100.49±6.92 | 0.23 |
| Mild | 39 | 28.17±9.71 |  | 156.32±12.12 |  | 99.23±7.48 |  |
| Moderate | 26 | 24.49±11.16 |  | 163.53±10.76 |  | 99.75±6.63 |  |
| Severe | 16 | 29.31±10.39 |  | 156.93±11.82 |  | 103.51±8.35 |  |

| L4/5 |  |  |  |  |  |  |  |
| --- | --- | --- | --- | --- | --- | --- | --- |
| Discopathy involvement: |  |  |  |  |  |  |  |
| Yes | 179 | 26.97±10.36 | 0.43 | 159.12±10.71 | 0.164 | 100.31±7.17 | 0.638 |
| No | 21 | 28.91±12.8 |  | 162.62±12.06 |  | 101.09±7.03 |  |
| Discopathy types: |  |  |  |  |  |  |  |
| No Discopathy | 21 | 28.91±12.8 | 0.764 | 162.62±12.06 | 0.476 | 101.09±7.03 | 0.692 |
| Disc protrusion | 10 | 23.61±7.92 |  | 162.26±10.15 |  | 98.15±6.58 |  |
| Disc herniation | 17 | 24.85±11.44 |  | 162.17±9.32 |  | 100.97±5.94 |  |
| Disc bulge | 140 | 27.44±10.11 |  | 158.52±10.9 |  | 100.58±7.37 |  |
| Disc cephalic migration | 3 | 28.07±10.3 |  | 158.67±11.38 |  | 96±8.42 |  |
| Disc caudal migration | 9 | 27.18±15.05 |  | 159.44±11.11 |  | 98.62±6.67 |  |
| Discopathy Severity: |  |  |  |  |  |  |  |
| Normal | 21 | 28.91±12.8 | 0.796 | 162.62±12.06 | 0.587 | 101.09±7.03 | 0.241 |
| Mild | 78 | 27.46±9.4 |  | 159.1±11.11 |  | 99.81±5.93 |  |
| Moderate | 72 | 26.82±10.66 |  | 159.15±10.48 |  | 99.87±8.13 |  |
| Severe | 29 | 26.04±12.22 |  | 159.12±10.5 |  | 102.73±7.42 |  |
| L5/S1 |  |  |  |  |  |  |  |
| Discopathy involvement: |  |  |  |  |  |  |  |
| Yes | 134 | 26.35±10.58 | 0.117 | 160.24±11.2 | 0.165 | 100.27±7.57 | 0.743 |
| No | 66 | 28.86±10.59 |  | 157.97±10.09 |  | 100.63±6.22 |  |
| Discopathy types: |  |  |  |  |  |  |  |
| No Discopathy | 66 | 28.86±10.59 | 0.135 | 157.97±10.09 | 0.199 | 100.63±6.22 | 0.829 |
| Disc protrusion | 19 | 26.56±8.46 |  | 157.68±10.22 |  | 100.01±6.76 |  |
| Disc herniation | 19 | 22.92±8.75 |  | 161.85±9.49 |  | 100.7±8.76 |  |
| Disc bulge | 90 | 26.54±11.24 |  | 160.6±11.44 |  | 100.42±7.67 |  |
| Disc cephalic migration | 2 | 41.9±9.48 |  | 144.35±17.32 |  | 97.1±4.53 |  |
| Disc caudal migration | 3 | 27.33±5.62 |  | 165.87±15.18 |  | 100.43±3.09 |  |
| Disc sequestration | 1 | 36.6±0.00 |  | 160.6±0.00 |  | 89.6±0.00 |  |
| Discopathy Severity: |  |  |  |  |  |  |  |
| **Normal** | 66 | 28.86±10.59 | 0.129 | 157.97±10.09 | 0.517 | 100.63±6.22 | 0.787 |
| **Mild** | 67 | 26.49±10.9 |  | 159.73±(11.22) |  | 99.67±7.71 |  |
| **Moderate** | 46 | 27.77±10.68 |  | 160.53±11.97 |  | 100.85±6.94 |  |
| **Severe** | 21 | 22.79±8.81 |  | 161.25±9.72 |  | 100.95±8.61 |  |

The analysis indicates no statistically significant relationship among LLA, SKA, and STA in discopathy involvement, types, and severity. However, the SKA shows significant variation with discopathy severity at L3/4 (*p*=0.044), particularly between the mild and moderate groups (*p*=0.009). Examples of morphological LPAs for the last three lumbosacral levels (L3/4, L4/5, and L5/S1), with disc severity classified as mild, moderate, and severe, are provided in [Supplementary S5]

### The Relationships of LPA Changes with Age, Gender, and BMI

The relationship between age classification across age subgroups (young, middle, and old), gender (male and female), and BMI classification (normal weight, overweight, and obese) are summarised in Table 5, and examples of LPA morphometric measures (LLA, SKA, and STA) based on age and BMI classification are illustrated in [Supplementary S6 and S7].

**Table 5.** Distribution of the relationship between lumbo-pelvic morphometric (LPA) changes and age, gender, and body mass index (BMI) classification (n=200). N: number of participants; SD: standard deviation; * relationship is significant

| Variables | Lumbo-Pelvic Morphometric |  |  |  |  |  |  |
| --- | --- | --- | --- | --- | --- | --- | --- |
|  | Lumbar Lordosis Angle |  |  | Sacral Kyphosis Angle |  | Sacral Table Angle |  |
|  | N | Mean±SD | *p* | Mean±SD | *p* | Mean±SD | *p* |
| Age Classification: |  |  |  |  |  |  |  |
| Young age | 73 | 26.64±10.04 | 0.522 | 158.9±11.39 | 0.237 | 99.77±6.63 | 0.523 |
| Middle age | 74 | 26.68±10.1 |  | 161.14±9.99 |  | 101.1±6.89 |  |
| Old age | 53 | 28.61±12.09 |  | 158.01±11.24 |  | 100.24±8.14 |  |
| Gender Classification: |  |  |  |  |  |  |  |
| Male | 105 | 24.01±9.14 | <0.001* | 160.05±9.96 | 0.443 | 99.12±6.54 | 0.008* |
| Female | 95 | 30.68±11.08 |  | 158.87±11.83 |  | 101.79±7.54 |  |
| BMI Classification: |  |  |  |  |  |  |  |
| Normal weight | 59 | 24.03±9.93 | 0.004* | 160.88±11.29 | 0.481 | 101.39±6.11 | 0.414 |
| Overweight | 57 | 26.47±10.13 |  | 158.53±11.32 |  | 99.7±7.4 |  |
| Obese | 84 | 29.87±10.85 |  | 159.17±10.29 |  | 100.15±7.63 |  |

Age classification showed no statistical significance for lumbo-sacral measurements or lumbo-pelvic morphometrics. Regarding gender, significant relationships were found: males had a lower mean LLA (24.01±9.14) than females (30.68±11.08, *p*<0.001), and similarly at STA (99.12±6.54 vs 101.79±7.54, *p*=0.008). BMI was significantly associated with LLA, increasing from normal weight (24.03±9.93) to overweight (26.47±10.13) and obese (29.87±10.85), with significant differences mainly between normal and obese groups (*p*=0.004).

### Set Standardisation of Local LPA Morphometric Measurements for Discopathic Patients

Correlation between LPA changes and LBP duration classification (Acute, Subacute, and Chronic), summarised in Table 6. Participants with acute LBP (<6 weeks) had mean LLA, SKA and STA of 27.28°±11.35°, 159.8°±10.19°, and 99.03°±7.58°, with no significant correlations. In the sub-acute group (6 to 12 weeks), these were 27.04°±10.96°, 162.97°±10.11°, and 100.4°±6.8°, with a moderate negative correlation between LLA and SKA (r= −0.592, *p*= 0.006). In the chronic group (≥12 weeks), the angles were 27.18°±10.53°, 158.99°±11.06°, and 100.62°±7.13°, with a similar negative correlation (r = −0.443, *p*<0.001).

**Table 6.**
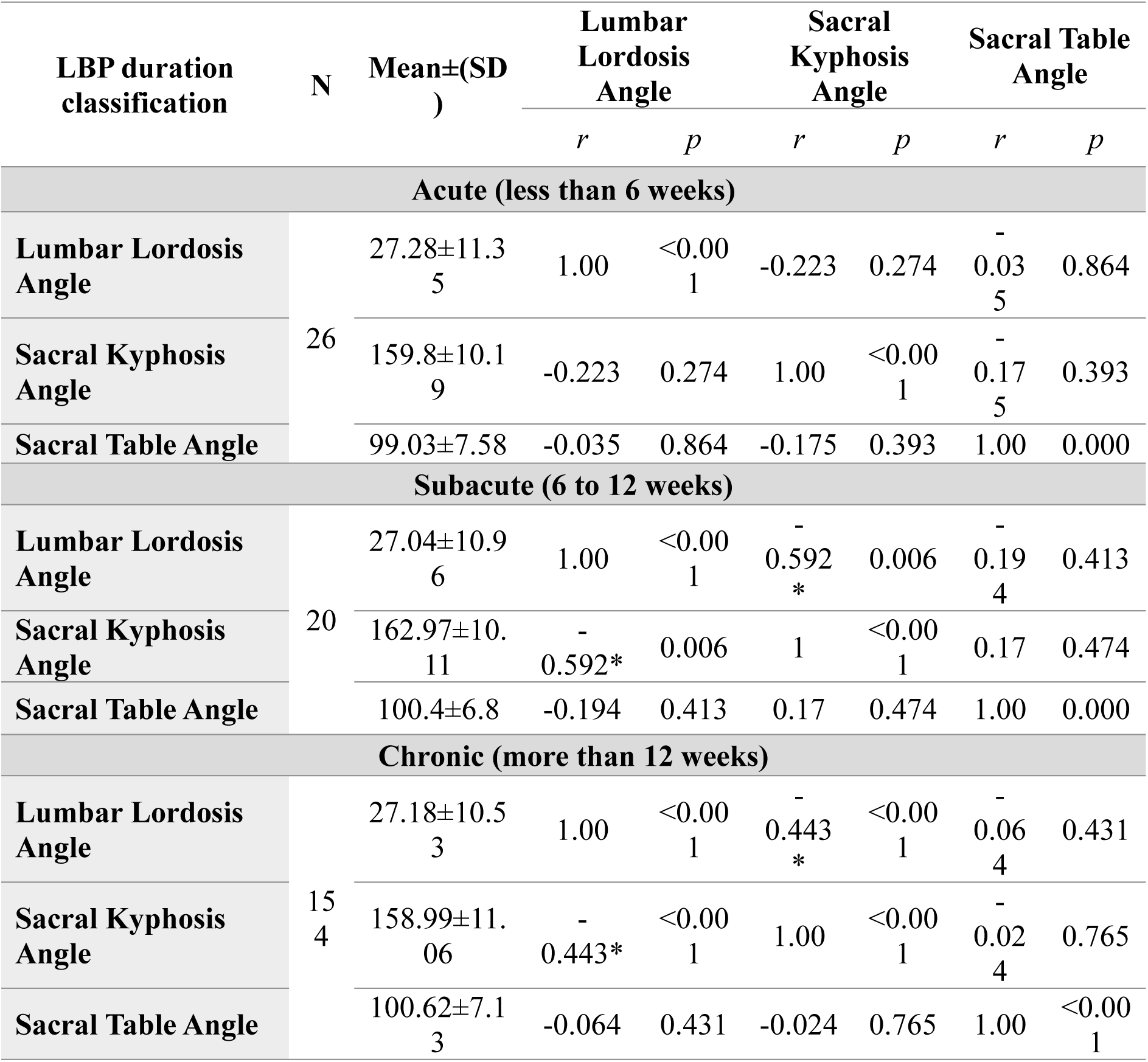
The set standardisation and correlation between lumbo-pelvic morphometric (LPA) changes and lower back pain (LBP) duration classification among discopathic patients (n=200). N: number of participants; SD: standard deviation; * Correlation is significant at the p<0.05

## Discussion

This study examined whether MRI-derived LPAs (LLA, SKA, and STA) are associated with discopathy characteristics and clinical burden in adults with low LBP referred for MRI in the Gaza Strip. The cohort predominantly comprised individuals with chronic symptoms and moderate-to-severe functional limitation, reflecting a clinically selected population typically referred for advanced imaging. MRI findings were mainly at lower lumbar levels, particularly at L4/5 and L5/S1, with disc bulges most common. Overall, LPAs demonstrated limited clinical relevance. No meaningful associations were observed between these morphometric measures and discopathy involvement, type, or severity, nor with disability as measured by ODI. A weak positive correlation was identified between STA and LBP duration, and an isolated association between SKA and discopathy severity was observed only at the L3/4 level. These findings suggest that static MRI-based lumbopelvic alignment parameters have limited value in explaining symptom severity or structural disc pathology, aligning with earlier evidence that shows weak links between structure and symptoms in LBP [34–36]

The cohort consists of a clinically selected group usually referred for MRI due to chronic symptoms, moderate-to-severe disability, and frequent radiating pain or neurological issues. This referral pattern is significant because MRI is typically used for persistent or complex LBP, which limits the generalizability of findings to the general population with non-specific LBP [37]. Furthermore, MRI findings showed that discopathy mainly affected L4/5 and L5/S1, with disc bulge the most common abnormality; caudal migration was more common than cephalic migration, and Modic type 2 changes were primarily seen at lower lumbar levels, whereas Schmorl’s nodes were more common at upper levels. These patterns align with established MRI research and reflect the biomechanical loading characteristics of the lumbosacral spine[10, 38–41].

Gender and BMI-related differences in lumbo-pelvic alignment were observed, with males exhibiting lower LLA and STA, while obese participants showed increased lumbar lordosis. These results align with previous studies that link differences in alignment to pelvic structure and compensatory responses [42–45]. However, these variations did not correlate with differences in pain duration or disability, indicating that static alignment measures may have limited clinical significance in discopathy-related lower back pain.

Despite the high prevalence of disc abnormalities, LPAs (LLA, SKA, STA) did not differ by discopathy involvement, type, or severity, indicating that these angles have limited value for discriminating discopathic phenotypes in symptomatic adults. This is consistent with Walter et al., who reported no significant associations between lumbar lordosis, sacral slope, pelvic tilt, or pelvic incidence and Pfirrmann-graded disc degeneration, and likewise found no relationship with back pain outcomes despite frequent degenerative changes [35]. Although sagittal parameters are strongly constrained by constitutional pelvic morphology and geometric coupling, meaning they often reflect innate alignment rather than disc pathology [46]. However, the isolated association of SKA severity with L3/4 was not replicated at adjacent levels. It should therefore be interpreted cautiously, particularly given multiple comparisons and the absence of a consistent pattern. Additionally, LPAs were mainly unrelated to clinical measures align with evidence that lordosis-related metrics are often weakly related to disability in chronic LBP, supporting multifactorial models in which symptoms extend beyond static structural or alignment measures [47].

Duration-stratified analysis of LPAs revealed typical internal spinal geometry across acute, subacute, and chronic LBP, with similar mean LLA, SKA, and STA values. Moderate negative correlations between LLA and SKA in subacute and chronic groups suggest consistent geometric coupling that isn’t linked to pain severity, disability, or discopathy. These patterns serve as descriptive references within a symptomatic cohort, not as indicators of disease progression. However, methodological factors probably influenced the limited correlations seen. Measurements obtained from supine MRI do not reflect sagittal alignment under normal loading conditions. Previous research has shown consistent differences between supine and standing alignment, especially at the lumbosacral junction, which may reduce the strength of links with functional outcomes [48, 49].

This study has several limitations that impact the interpretation and generalisability of its findings. Its cross-sectional design precludes causal inference and limits assessment of temporal relationships between lumbo-pelvic morphology and discopathy. Furthermore, the design cannot determine whether specific LPA patterns predispose to discopathy or result from disc degeneration. Data collection during the COVID-19 pandemic may have introduced bias due to lower referral rates and the absence of asymptomatic controls, limiting the ability to differentiate morphological features between symptomatic and asymptomatic cases. The use of different MRI scanners without a formal assessment of intra- or inter-observer reliability further introduces potential measurement variability. Subgroup analyses were limited by sample size, and multiple comparisons increased the risk of type I error. Finally, the absence of asymptomatic controls restricts generalisability and interpretation of the proposed reference values.

## Conclusion

MRI enables reproducible assessment of LPA; however, these measures showed limited association with discopathy features pain duration, or disability in adults with LBP. Apart from a weak association between STA and symptom duration and an isolated level-specific finding for SKA, LPA did not meaningfully explain clinical or structural variability. Importantly, this study establishes population-specific reference values for LPA (LLA, SKA and STA) in adults with discopathy-related LBP in the Gaza Strip. These reference values provide benchmarks for this population and should be interpreted cautiously, within a broader clinical and biomechanical framework rather than as standalone indicators of disease severity.

## Contribution Statement

AM was responsible for data collection, image analysis and interpretation. YA and AN contributed to study conception and design and critically revised the manuscript for important intellectual content. FA contributed to data interpretation and drafting of the manuscript. KM contributed to the analysis and interpretation of data and led manuscript drafting and critical revision for scientific rigour. All authors approved the final version of the manuscript and agree to be accountable for all aspects of the work

## Ethics declarations

Ethical approval was granted by the Helsinki Committee (HC) for Ethical Approval, Palestinian Health Research Council (PHRC), Gaza Strip, Palestine (approval number PHRC/HC/713/20; 01 June 2020). Administrative approval was obtained from the Ministry of Health, State of Palestine (Human Resources Development Directorate; correspondence number 504832; 16 June 2020). Academic approval was granted by the Faculty of Applied Medical Sciences, Al-Azhar University-Gaza (03 June 2020). Written informed consent was obtained from all participants prior to participation.

## Informed Consent Statement

All participants provided written informed consent before participation.

## Conflict of interest

The authors declare that they have no conflicts of interest.

## Funding

The authors declare that no funding was received for this study.

## Data availability

Data supporting the findings of this study are included in the article and Supplementary Materials; further data are available from the corresponding author upon reasonable request.

## Notes

### Competing Interest Statement

The authors have declared no competing interest.

### Funding Statement

This study did not receive any funding.

