## Supplementary materials for "MRI-derived lumbo-pelvic angles and lumbar discopathy in adults with low back pain from the Gaza Strip: a cross-sectional study"

**S1: Pilot Study**

This was conducted before data collection began to identify issues in the research design and to test the validity and reliability of the data collection tools [MRI scanners, RadiAnt Viewer, and the questionnaire]. According to the American College of Radiology [1], the geometric accuracy quality control test uses an MRI water phantom with known real length and diameter to ensure validity across both MRI scanners at the EGH and Al-Shifa hospitals. A phantom with actual dimensions of 29.5 cm in length and 13.5 cm in diameter was used. Both MRI scanners imaged the phantom with a spine coil; the results were compared with the real measurements. MRI phantom image measurements were within ±2 mm of their actual values, which conform to the ACR-recommended action criteria for measured lengths and diameters. [Figures 1 and 2 illustrate the validity of MRI scanners and RadiAnt DICOM]. Additionally, the intra-rater reliability of MRI scanners and the RadiAnt DICOM Viewer was assessed at EGH and Al-Shifa Hospital. One investigator took ten measurements of length and diameter on MRI phantom images. Means were calculated in SPSS, and the ICCs were 0.90 for length and 0.92 for diameter, indicating excellent intra-rater reliability. To confirm Radiant Software's validity and reliability in measuring angles, a water phantom with known dimensions (length = 13.5 cm, width = 29.5 cm) was tested. It was imaged on a coronal T2-WI, and measurements were taken using a Radiant reader programme. The angles were calculated mathematically via the Pythagorean theorem (https://www.calculator.net/right-triangle-calculator.html) and compared with those from the program. The results showed excellent agreement, indicating the tool's accuracy for angle measurement Figure 3.

Furthermore, to ensure the questionnaire's validity, the researcher submitted it to an expert panel [Table 1] with experience and knowledge of the topic, which provided suggestions and judgments on the instrument's adequacy. To assess the questionnaire's reliability, Cronbach's alpha was calculated in SPSS, measuring reliability within each domain and across the entire instrument. The researcher estimated the instrument's reliability using Cronbach’s alpha (34 items), yielding an alpha of 0.716, indicating high reliability. The normal range of Cronbach’s alpha is between 0.00 and 1.0, and the interclass correlation coefficient was 0.004 (highly significant).

Finally, according to Fisher and Johnston [2], the validation of the Oswestry Disability Questionnaire, its sensitivity as a measure of change following treatment, and its relationship with other aspects of the chronic pain experience, physiotherapy theory, and practice are discussed. The reliability of the ODI addresses a broader concept of disability than simply pain intensity [3]. The internal consistency of the Yoruba ODI, measured with Cronbach's alpha, was 0.97; intra-rater reliability was assessed using an intra-class correlation coefficient of 0.93; and criterion validity was evaluated through Spearman's rank correlation, resulting in r = 0.92 for the highest score and 0.63 for the lowest score [4]. Overall, the ODI has been extensively tested for validity and reliability across various settings, as reported in several studies[5-8].

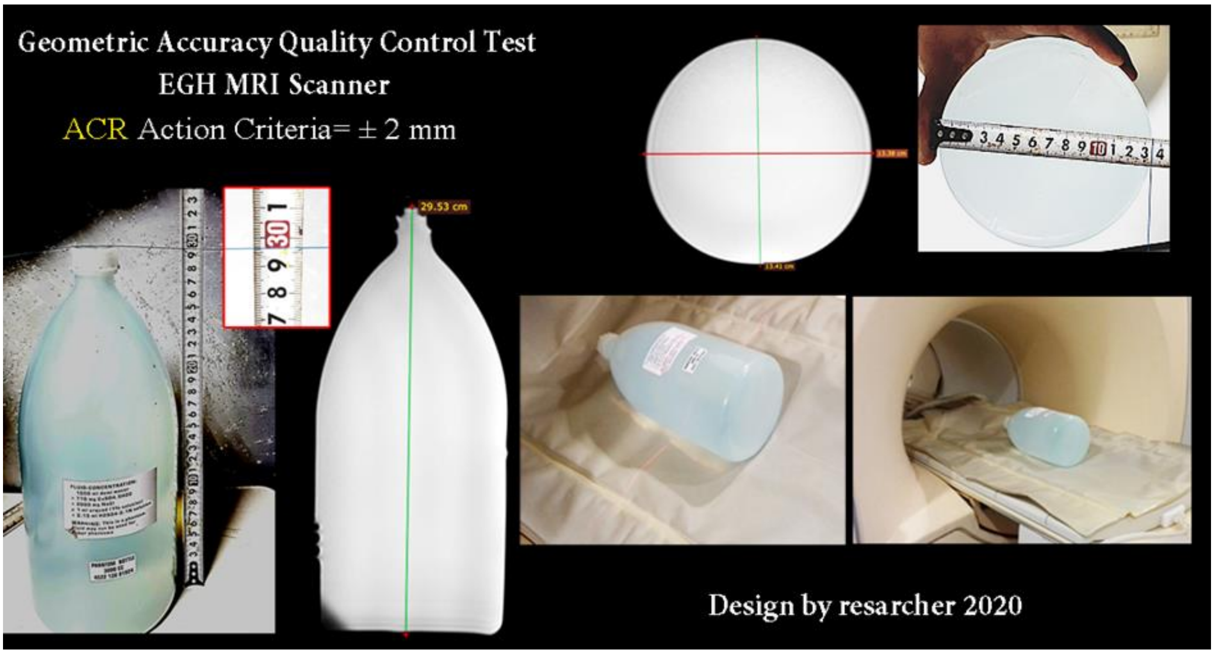


*Figure 1: Validity EGH MRI scanner.*

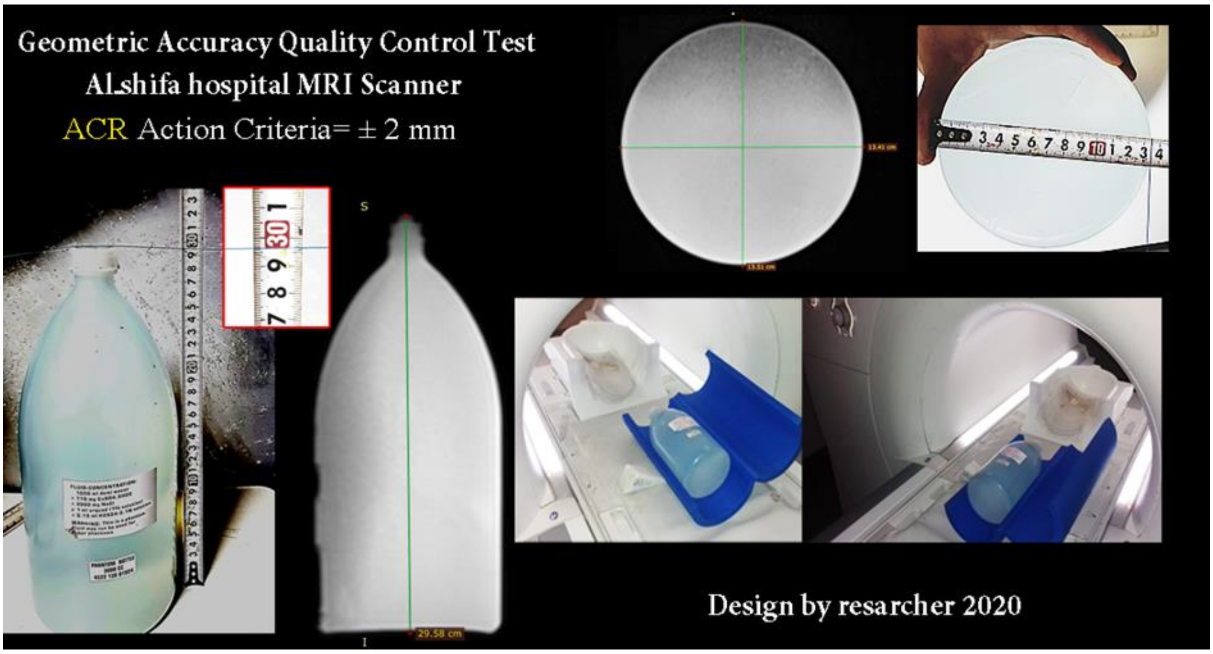


*Figure 2. Validity Al-Shifa hospitals MRI scanner.*

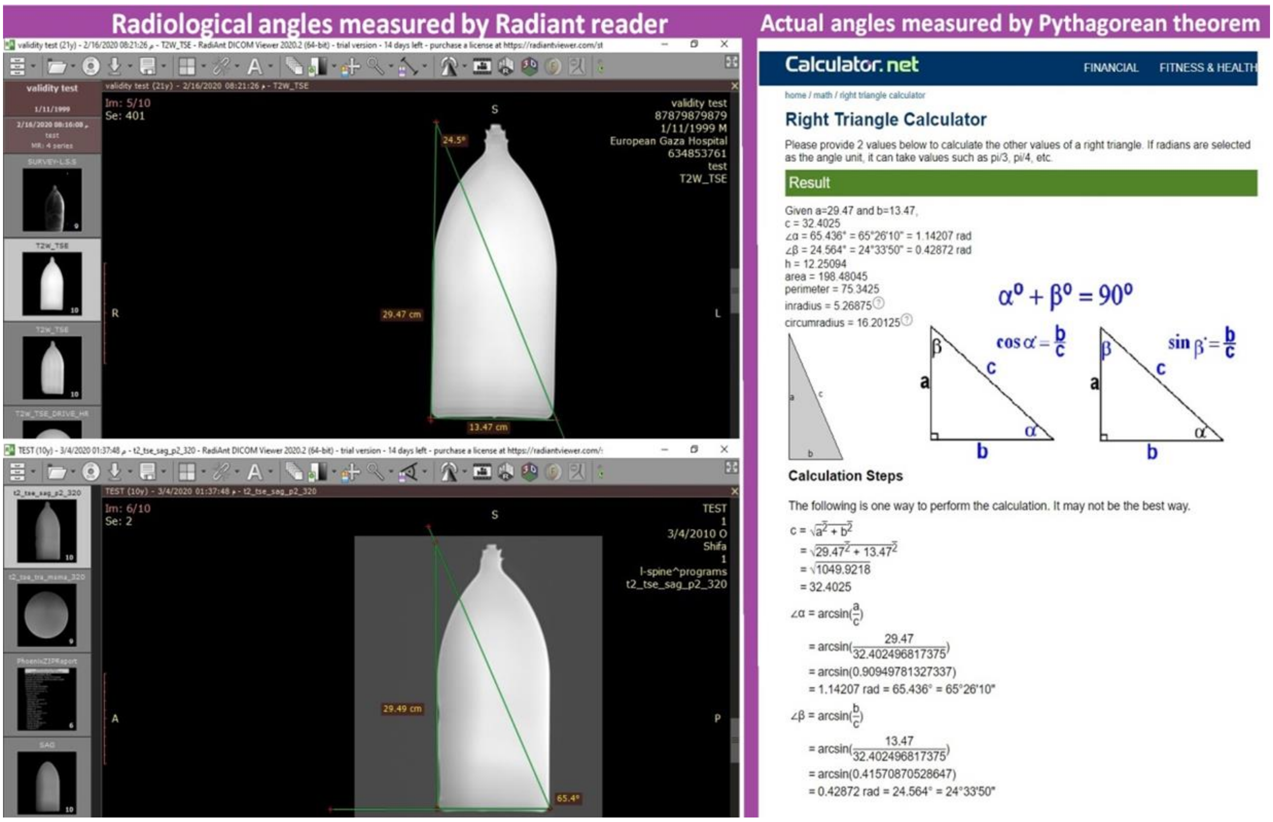


*Figure 3: Radiological angles measured by Radiant reader.*

*Table 1. Expert Panel List of arbitrators from radiologists, neurosurgeons, orthopedists, and physiotherapists.*

| Name | Affiliation |
|---|---|
| **Dr Sadi Jaber** | Radiology Consultant, Nasser Medical Complex |
| **Dr Samy Al Agha** | Assistant Professor, Al Azhar University - Gaza |
| **Dr Samer Hamad** | Radiology Consultant, European Gaza Hospital |
| **Dr Marwan Matar** | Radiology Consultant, European Gaza Hospital |
| **Dr Hassan Abdul-Rahim** | Radiology Consultant, Al Khafji Joint Operation Hospital |
| **Dr Abdalrahem Mousa** | Head of Physiotherapy, Nasser Medical Complex |
| **Dr Nedal Abo Hadrous** | Head of Neurosurgery, European Gaza Hospital |
| **Dr Mohab Mousa** | Neurosurgery Specialist, Kharki Medical Academy Postgraduate Education |
| **Dr Jamal Abo Helal** | Orthopaedic Consultant, European Gaza Hospital |
| **Dr Mouhmad Al Qaraa** | Orthopaedic Consultant, European Gaza Hospital |
| **Mr Hussam Mansour** | Assistant Lecturer, Al Azhar University - Gaza |

**S2: Radiological diagnostic finding**

### Level L1/2

**Discopathy**: Yes ( ) No ( )

**Discopathy Types :**

No ( ) Disc protrusion ( ) Disc herniation ( ) Disc bulge ( )

Disc cephalic migration **( )** **Disc caudal migration ( )** **Disc sequestration ( )**

**Schmorl's nodes** Yes ( ) No ( )

**Schmorl's nodes side** 1.No 2. centre 3. peripheral

**Discopathy affected side :**

No ( ) Right side ( ) central ( ) left side ( ) Both sides ( )

**Discopathy location :**

No ( ) Central zone ( ) Sub articular zone ( ) foramina( ) Extra-foramina ( )

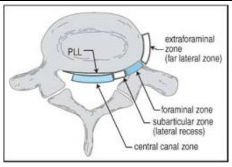


**Discopathy Severity :** Normal ( ) mild ( ) moderate ( ) severe ( )

**Spondylotic changes**: Yes ( ) No ( )

**Spinal canal stenosis:** Yes ( ) No ( ) , If yes, how the measurement = **-------------**

If yes, how? , Right ( ) or Left ( ) Both sides ( )

**Flavum ligament hypertrophy:** Yes ( ) No ( ) , If yes, how the measurement = **---------**

If yes, how? Right ( ) or Left ( ) Both sides ( )

**Facet joint arthritis :** Yes ( ) No ( ) , If yes, how Right ( ) Left ( ) Both sides ( )

**Spondylolesthesis:** Yes ( ) No ( ) **,** If yes, what is the grade: 1 ( ) 2 ( ) 3 ( ) 4 ( )

**Retrolisthesis:** Yes ( ) No ( ) If yes, what is the grade: 1 ( ) 2 ( ) 3 ( ) 4 ( )

**Modic changes endplate** : Yes ( ) No ( ) If yes, what is the grade: 1 ( ) 2 ( ) 3 ( )

### Level L2/3

**Discopathy**: Yes ( ) No ( )

**Discopathy Types :**

No ( ) Disc protrusion ( ) Disc herniation ( ) Disc bulge ( )

Disc cephalic migration **( )** **Disc caudal migration ( )** **Disc sequestration ( )**

**Schmorl's nodes** Yes ( ) No ( )

**Schmorl's nodes side** 1.No 2. centre 3. peripheral

**Discopathy affected side :**

No ( ) Right side ( ) central ( ) left side ( ) Both sides ( )

**Discopathy location :**

No ( ) Central zone ( ) Sub articular zone ( ) foramina( ) Extra-foramina ( )

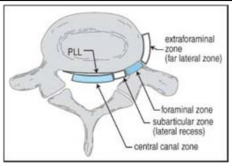


**Discopathy Severity :** Normal ( ) mild ( ) moderate ( ) severe ( )

**Spondylotic changes**: Yes ( ) No ( )

**Spinal canal stenosis:** Yes ( ) No ( ) If yes, how the measurement = **-------------**

If yes, how? Right ( ) or Left ( ) Both sides ( )

**Flavum ligament hypertrophy:** Yes ( ) No ( ) If yes, how the measurement = **---------**

If yes, how? Right ( ) or Left ( ) Both sides ( )

**Facet joint arthritis :** Yes ( ) No ( ) If yes, how Right ( ) Left ( ) Both sides ( )

**Spondylolesthesis:** Yes ( ) No ( ) If yes, what is the grade: 1 ( ) 2 ( ) 3 ( ) 4 ( )

**Retrolisthesis:** Yes ( ) No ( ) If yes, what is the grade: 1 ( ) 2 ( ) 3 ( ) 4 ( )

**Modic changes endplate** : Yes ( ) No ( ) If yes, what is the grade: 1 ( ) 2 ( ) 3 ( )

### Level L3/4

**Discopathy**: Yes ( ) No ( )

**Discopathy Types :**

No ( ) Disc protrusion ( ) Disc herniation ( ) Disc bulge ( )

Disc cephalic migration ( ) Disc caudal migration ( ) Disc sequestration ( )

**Schmorl's nodes** Yes ( ) No ( )

**Schmorl's nodes side** 1.No 2. Centre 3. peripheral

**Discopathy affected side :**

No ( ) Right side ( ) Central ( ) Left side ( ) Both sides ( )

**Discopathy location :**

No ( ) Central zone ( ) Subarticular zone ( ) Foramina( ) Extra-foramina ( )

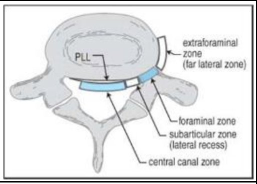


**Discopathy Severity :** Normal ( ) **M**ild ( ) Moderate ( ) Severe ( )

**Spondylotic changes**: Yes ( ) No ( )

**Spinal canal stenosis:** Yes ( ) No ( ) If yes, how the measurement = **-------------**

If yes, how? Right ( ) or Left ( ) Both sides ( )

**Flavum ligament hypertrophy:** Yes ( ) No ( ) If yes, how the measurement = **---------**

If yes, how? Right ( ) or Left ( ) Both sides ( )

**Facet joint arthritis :** Yes ( ) No ( ) If yes, how Right ( ) Left ( ) Both sides ( )

**Spondylolesthesis:** Yes ( ) No ( ) If yes, what is the grade: 1 ( ) 2 ( ) 3 ( ) 4 ( )

**Retrolisthesis:** Yes ( ) No ( ) If yes, what is the grade: 1 ( ) 2 ( ) 3 ( ) 4 ( )

**Modic changes endplate** : Yes ( ) No ( ) If yes, what is the grade: 1 ( ) 2 ( ) 3 ( )

### Level L4/5

**Discopathy**: Yes ( ) No ( )

**Discopathy Types :**

No ( ) Disc protrusion ( ) Disc herniation ( ) Disc bulge ( )

Disc cephalic migration **( )** **Disc caudal migration ( )** **Disc sequestration ( )**

**Schmorl's nodes** Yes ( ) No ( )

**Schmorl's nodes side** 1.No 2. Centre 3. Peripheral

**Discopathy affected side :**

No ( ) Right side ( ) Central ( ) Left side ( ) Both sides ( )

**Discopathy location :**

No ( ) Central zone ( ) Subarticular zone ( ) Foramina( ) Extra-foramina ( )

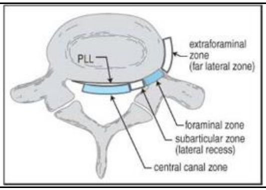


**Discopathy Severity :** Normal ( ) **M**ild ( ) Moderate ( ) Severe ( )

**Spondylotic changes**: Yes ( ) No ( )

**Spinal canal stenosis:** Yes ( ) No ( ) If yes, how the measurement = **-------------**

If yes, how? Right ( ) or Left ( ) Both sides ( )

**Flavum ligament hypertrophy:** Yes ( ) No ( ) If yes, how the measurement = **---------**

If yes, how? Right ( ) or Left ( ) Both sides ( )

**Facet joint arthritis :** Yes ( ) No ( ) If yes, how Right ( ) Left ( ) Both sides ( )

**Spondylolesthesis:** Yes ( ) No ( ) If yes, how the grade: 1 ( ) 2 ( ) 3 ( ) 4 ( )

**Retrolisthesis:** Yes ( ) No ( ) If yes, what is the grade: 1 ( ) 2 ( ) 3 ( ) 4 ( )

**Modic changes endplate** : Yes ( ) No ( ) If yes, what is the grade: 1 ( ) 2 ( ) 3 ( )

### Level L5/S1

**Discopathy**: Yes (   )                No (   )

**Discopathy Types :**

No (   )     Disc protrusion (   )     Disc herniation (   )     Disc bulge (   )

Disc cephalic migration **(   )**     **Disc caudal migration (   )**     **Disc sequestration (   )**

| | |
|---|---|
| **Schmorl's nodes** Yes ( ) No ( ) | **Schmorl's nodes side** 1.No 2. Centre 3. Peripheral |

**Discopathy affected side :**

No (   )     Right side (   )     Central (   )     Left side (   )     Both sides (   )

**Discopathy location :**

No (   )   Central zone (   ) Subarticular zone (   )   Foramina (   )   Extra-foramina (   )

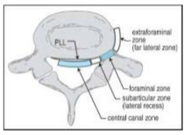


**Discopathy Severity :**   Normal (   )   Mild (   )   Moderate (   ) Severe (   )

| | |
|---|---|
| **Spondylotic changes**: Yes ( ) No ( ) | **Sacroiliitis**: Yes ( ) No ( ) If yes, how? Right ( ) or Left ( ) Both sides ( ) |

**Spinal canal stenosis:** Yes (   )   No (   ) If yes, how the measurement = **-------------**

If yes, how? Right (   ) or Left (   ) Both sides (   )

**Flavum ligament hypertrophy:** Yes (   )   No (   ) If yes, how the measurement = **---------**

If yes, how? Right (   ) or Left (   ) Both sides (   )

**Facet joint arthritis :** Yes (   )   No (   ) If yes, how Right (   )   Left (   )   Both sides (   )

**Spondylolesthesis:** Yes (   )   No (   ) If yes, what is the grade: 1 (   ) 2 (   ) 3 (   ) 4 (   )

**Retrolisthesis:** Yes (   )   No (   ) If yes, how the grade: 1 (   ) 2 (   ) 3 (   ) 4 (   )

**Modic changes endplate** : Yes (   )   No (   ) If yes, how the grade: 1 (   ) 2 (   ) 3 (   )

### S3: Distribution of the LBP duration classification in the study participants.

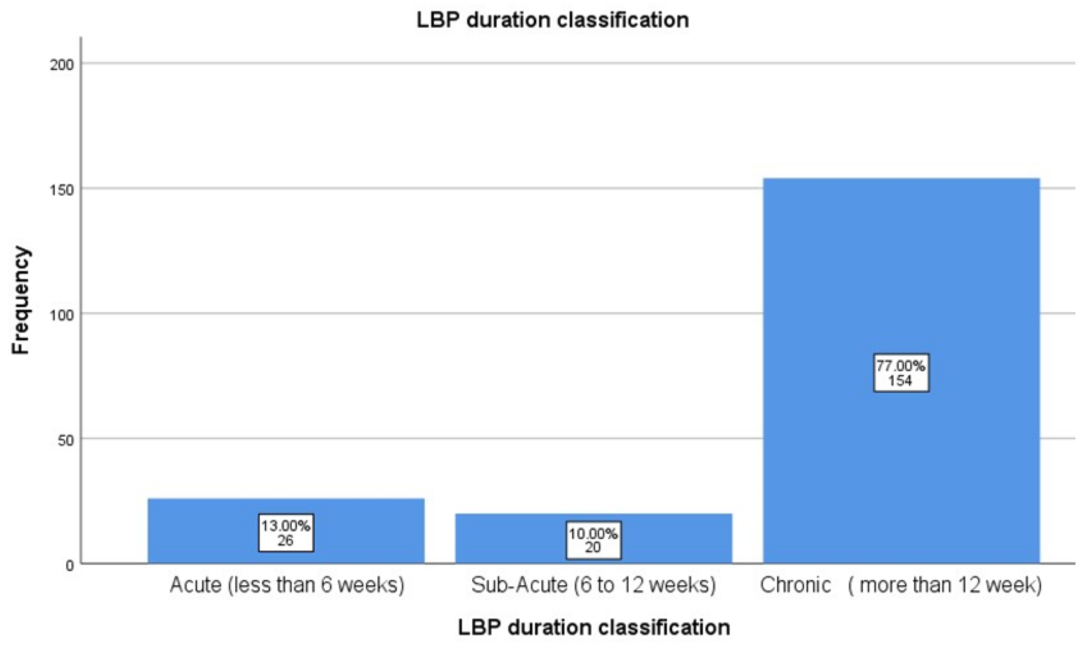


*Figure 4. Distribution of the LBP duration classification in the study participants.*

### S4: Distribution of the LBP disability classification in the study participants

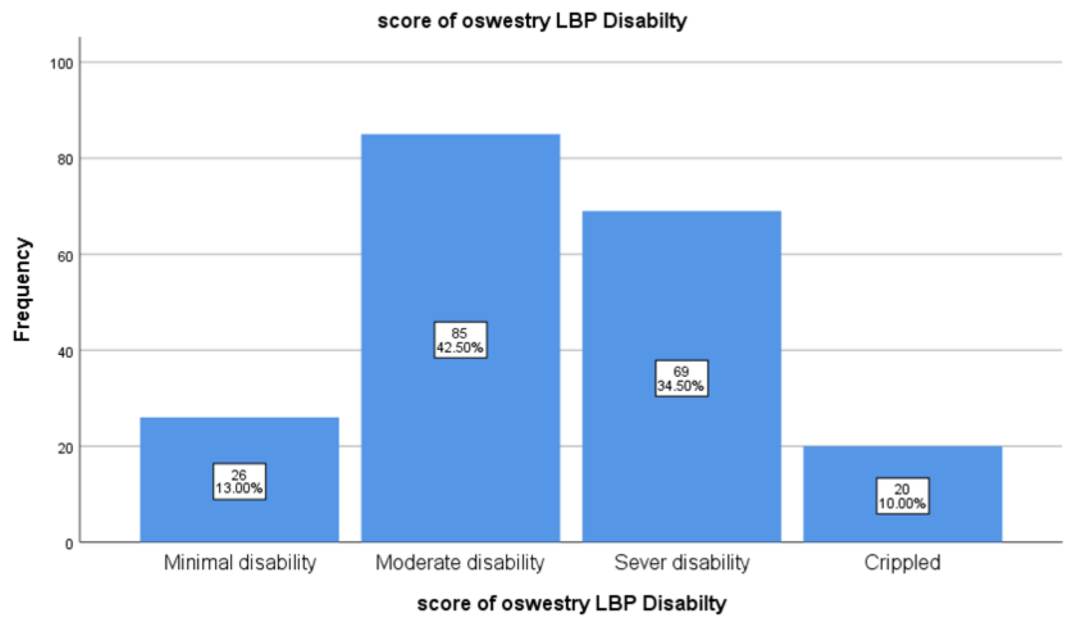


*Figure 5. Distribution of the LBP disability classification in the study participants.*

### S5: Examples of morphological lumbo-pelvic angles when the disc is mild, moderate, and severe at the last three lumbosacral levels.

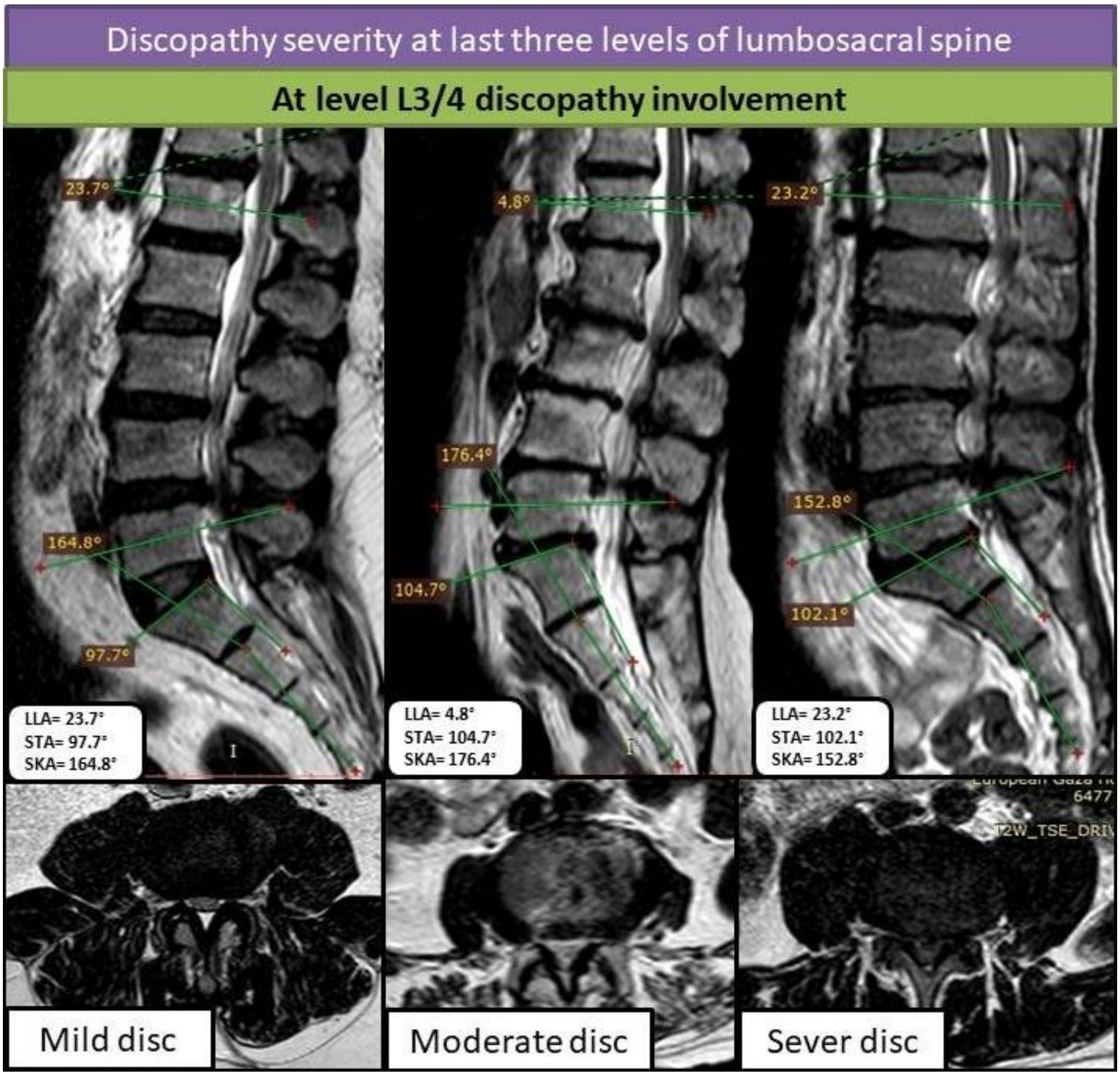


*Figure 6: The lumbo-pelvic angles within the discopathy: mild, moderate, and severe at the level L3/4 in axial and sagittal T2-WI*

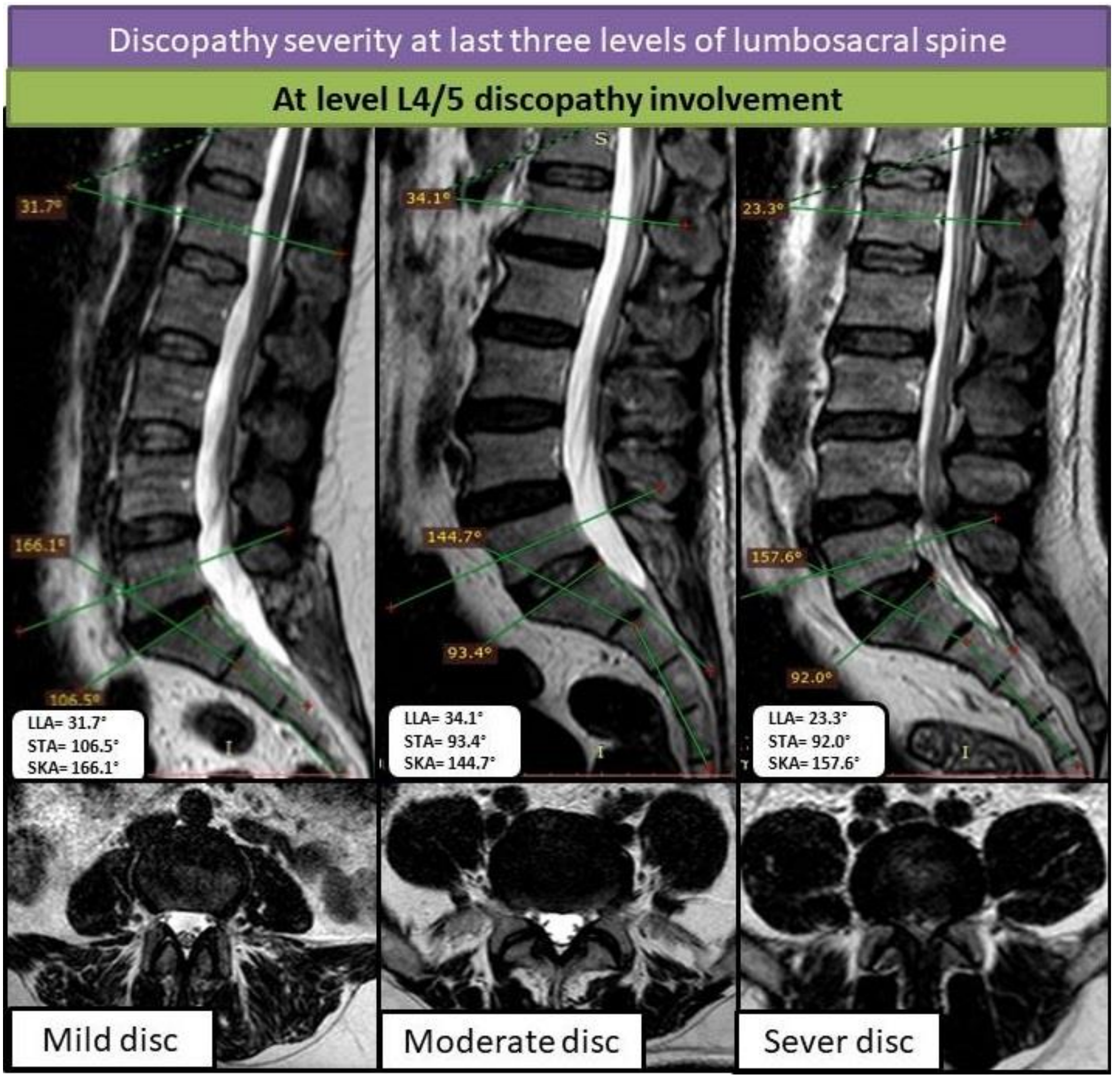


*Figure 7. The lumbo-pelvic angles within the discopathy: mild, moderate, and severe at the level L4/5 in axial and sagittal T2-WI.*

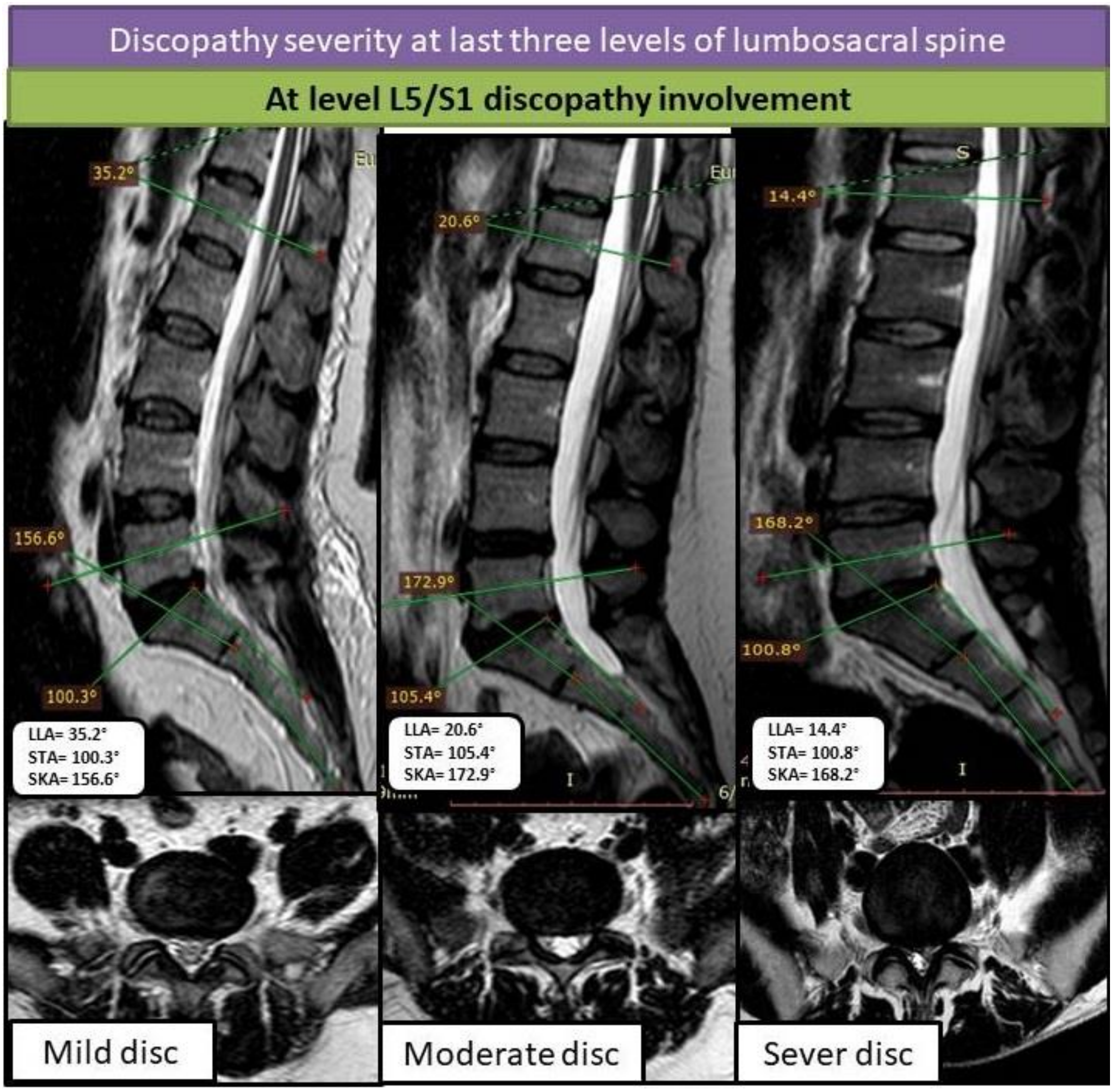


*Figure 8. displays the lumbo-pelvic angles within the discopathy: mild, moderate, and severe at the level L5/S1 in axial and sagittal T2-WI.*

**S6: Example of lumbo-pelvic morphometric measures (LLA, SKA, and STA)**

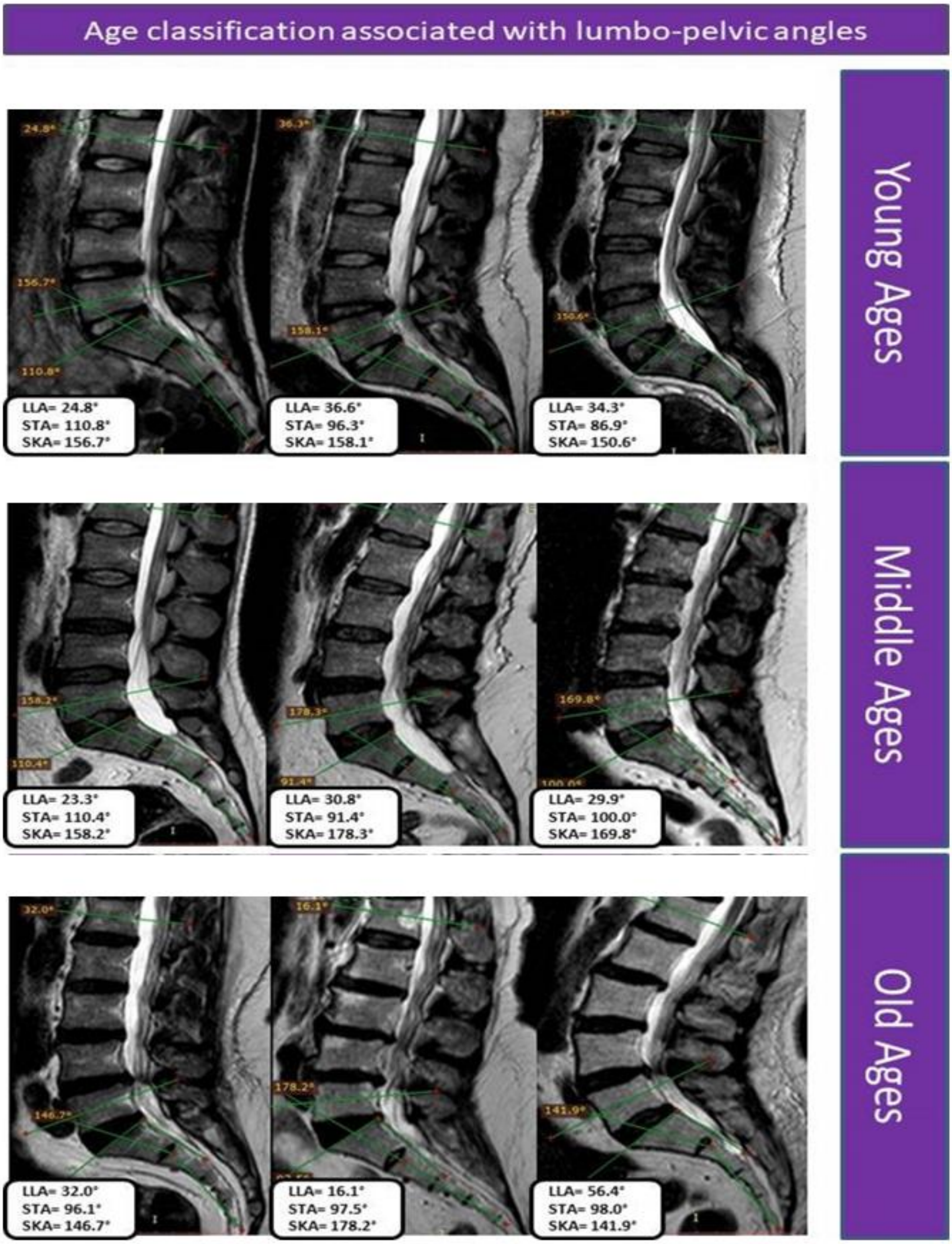


*Figure 9.Examples of morphological lumbo-pelvic angles with age sub-group in sagittal T2-WI.*

**S7: The examples of morphological lumbo-pelvic angles with BMI subgroups as (normal weight, overweight and obese) in sagittal T2-WI**

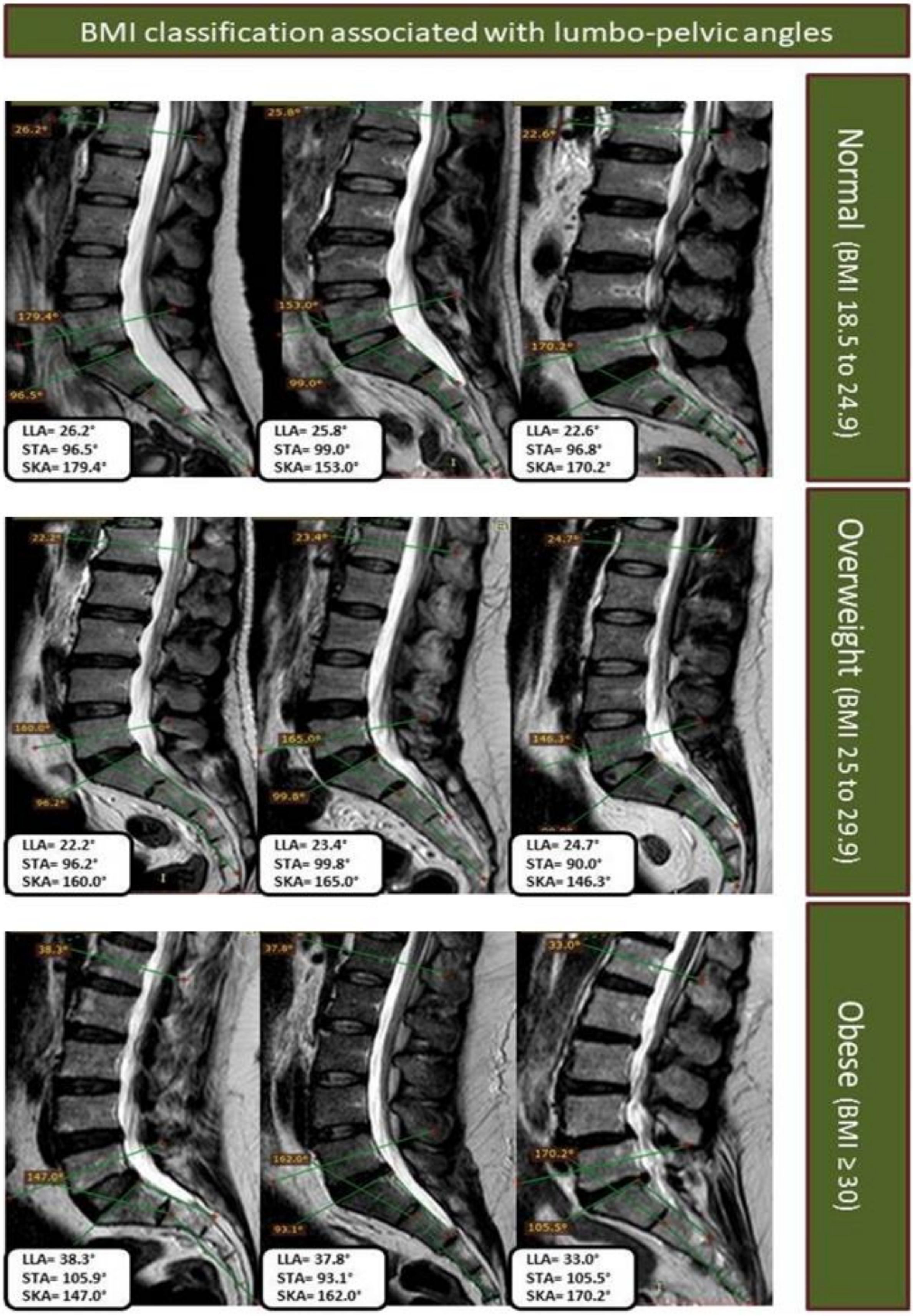

*Figure 10. The examples of morphological lumbo-pelvic angles with BMI subgroups as (normal weight, overweight and obese) in sagittal T2-WI*
